# Association between Cardiovascular Disease Risk and long COVID-19: A Systematic Review and Meta-analysis

**DOI:** 10.1101/2023.12.30.23300656

**Authors:** Yunxia Huang, Zhuofeng Wen, Hongbin Luo, Yijie Liao, Zisheng Chen

## Abstract

**Objectivate:** To assess demographic characteristics and investigate the correlation between cardiovascular diseases and long COVID-19 patients.

**Methods:** Comprehensive details, encompassing the first author’s name, publication year, sample inclusion criteria, sample size, and demographic characteristics, including age and gender of participants, were systematically extracted from all incorporated studies. Meanwhile, the analysis encompassed the assessment of the risk associated with nineteen cardiovascular outcomes.

**Results:** A total of 3, 201 potentially eligible studies were initially identified for consideration. Following rigorous literature screening and quality control measures, eighteen studies, encompassing 46, 083, 975 patients, met the specified criteria. In comparison to the control cohort, our investigation unveiled a heightened risk of 19 cardiovascular outcomes associated with long COVID-19. Our meta-analysis revealed a pooled OR of 1.68 (95% CI 1.55-1.81) (I^2^ = 69.1%, p= 0.000) for the overall risk of cardiovascular outcomes, indicating an elevated risk of cardiovascular diseases in individuals affected by long COVID-19. While the heterogeneity was relatively high, it is essential to acknowledge that all studies inherently carry an unavoidable risk of bias, irrespective of their quality rating. Moreover, there appears to be no significant publication bias based on the funnel plot, Begg’s, and Egger’s tests. Sensitivity analysis did not reveal substantial alterations in the overall stability of the results.

**Conclusions:** Our meta-analysis substantiates that individuals afflicted with long COVID-19 face an elevated risk of developing cardiovascular diseases. Routine assessment of cardiac function is deemed essential for a long COVID-19 high-risk cardiovascular disease population. Timely interventions have the potential to mitigate the risk of cardiovascular diseases and associated mortality in this population.

**TRIAL REGISTRATION PROSPERO Identifier:** CRD42023455701

## 1. Introduction

Since the COVID-19 global pandemic, there have been over 771, 151, 224 cumulative cases and 6, 960, 783 deaths of COVID-19 all over the world, as of 4 October 2023^1^. Recently, more and more researches have posited that COVID-19 may exert deleterious effects on patients beyond the acute phase, including potential negative impacts on respiratory, neurologic, mental, gastrointestinal, and cardiovascular health^2–5^. Consequently, the terminology ’long COVID’ or ’post-COVID-19 syndrome’ has emerged, specifically denoting the persistent clinical manifestations observed beyond the acute phase^6^. This fluctuating condition can be classified into two stages, namely post-acute and chronic, delineated by the duration of follow-up during which the symptoms manifest^7^. In the view of some studies, approximately one-eighth of the global population is projected to experience post-COVID-19 conditions. This implies that, considering over 651 million documented COVID-19 cases worldwide and a conservative estimate of a 10% incidence rate among infected individuals, around 65 million individuals worldwide may be affected by long COVID^8,9^. During the trajectory of long COVID-19, prevalent symptoms and signs include olfactory dysfunction, dyspnea, gustatory dysfunction, cough, fatigue, myalgia, among others, contributing to a diminished quality of life and decreased work efficiency^10^. Moreover, COVID-19 is implicated in significant multiorgan dysfunction, encompassing myocardial injury, acute respiratory distress syndrome, systemic shock, liver injury, renal injury, etc^11,12^.

Relevant studies suggest that the zinc metallopeptidase angiotensin-converting enzyme 2 (ACE2), responsible for converting angiotensin II to angiotensin, serves as the functional receptor for Severe Acute Respiratory Syndrome Coronavirus 2 CoV (SARS-CoV-2), whose interaction is implicated in myocardial damage, representing one potential mechanism, but the specific mechanisms remain elusive^13,14^. As matters stand, there is an increasing number of reports highlighting cardiovascular complications in long COVID, including arrhythmia, inflammatory heart disease, heart failure and various other cardiac disorders^15,16^. Actually, we have to acknowledge the existing controversy still surrounding the relationship between cardiovascular disease and long COVID-19. Specifically, regarding ischemic heart disease, findings from Hannah R Whittaker et al^2^ suggest a lower risk for long COVID-19 patients, while studies by Kwan Hong et al^17^ and Yan Xie et al^18^ present contrasting perspectives. Consequently, our current understanding of the association between cardiovascular disease and long COVID-19 remains limited. Further comprehensive evidence is imperative to establish a definitive association between cardiovascular disease and patients with long COVID-19.

As a consequence, we analyzed and reviewed relevant cohort studies to sort out the association of long COVID-19 patients with cardiovascular disease risk by the mean of meta-analysis comprehensively and deeply.

## 2. Materials and methods

### 2.1. Literature search

We performed a comprehensive meta-analysis of prospective and retrospective cohort studies documenting at least one combination of risk factors and outcomes. Our search strategy encompassed multiple databases, such as Web of Science, Scopus, Cochrane Library, Embase and PubMed, up to August 18, 2023, and was limited to articles published solely in English. Our search used the keywords "long COVID-19", "cardiovascular disease" and their Medical Subject Headings (MeSH) terms. Studies were required to fulfill the following medical standards: (1) only include prospective/retrospective cohort studies and case-control studies; (2) Patients with long COVID-19 were studied; (3) Outcome indicators: the risk of cardiovascular disease among patients with confirmed long COVID-19 (HR, RR and OR, with 95% CI); (4) Written in English only. Studies were excluded if they lacked the provision of OR/RR/HR with 95% CI, exhibited poor methodological quality, reported individual cases, or lacked the specified indicators of accomplishment mentioned above. Our research was registered in the Prospective Register of Systematic Reviews (PROSPERO ID CRD42023455701).

### 2.2. Data acquisition

Four investigators (Y. H., Z. W., H. L., Y. L.) independently extracted data and resolved discrepancies through consensus by four reviewers. The results underwent review by the senior investigator (Z. C.). When available, information on target outcomes was documented using WPS software. General details, including the first author’s name, publication year, sample inclusion criteria, sample size, and demographic data such as age and gender of study participants, were systematically collected from all included studies. Incidence outcomes were compared between the COVID-19 and non-COVID-19 groups.

### 2.3. Quality assessment

The risk of bias in the included cohort studies was evaluated using the Newcastle-Ottawa Scale (NOS). Studies achieving a score of at least 7 out of 10 were classified as high-quality studies.

### 2.4. Statistical analyses in Meta-analysis

We conducted meta-analyses using a statistical software package (Stata 11.0). Heterogeneity among the studies was assessed through χ2 analysis. If the inconsistency values (I^2^) exceeded 50%, a REM was applied; otherwise, a fixed-effect model (FEM) was utilized. In the presence of heterogeneity, we explored its source through subgroup analysis and sensitivity analysis. The combined HR, RR, OR, with its corresponding 95% CI was computed. Subsequently, funnel plots, Begg and Egger bias index tests, and meta-regression were employed to identify potential publication bias. All P values were two-sided, and statistical significance was set at P < 0. 05.

## 3. Results

### 3.1. Search results and study characteristics

The selection process is depicted in Figure. 1 via a flowchart. 3, 201 studies were deemed potentially eligible across various databases. 591 duplicates were excluded, then 2610 left. After extensively comparing the studies’ relevant information, 1, 001 researchers were included, but 1,609 were excluded (150 case reports, 371 conference abstracts, 197 comments, 20 editorials, 71 letters, 117 notes, 631 review, 52 meta-analyses). According to the studies for the title and abstract screening, 136 articles were taken into the next round, and 865 were dismissed (60 not related to COVID-19, 270 irrelevant research, 513 about other diseases, 22 not cohort studies). Finally, eighteen articles remained after reading the full articles, and 118 were ruled out (17 studies with incomplete outcomes, 88 irrelevant researches, 13 irrelevant interventions).

**Figure 1.**
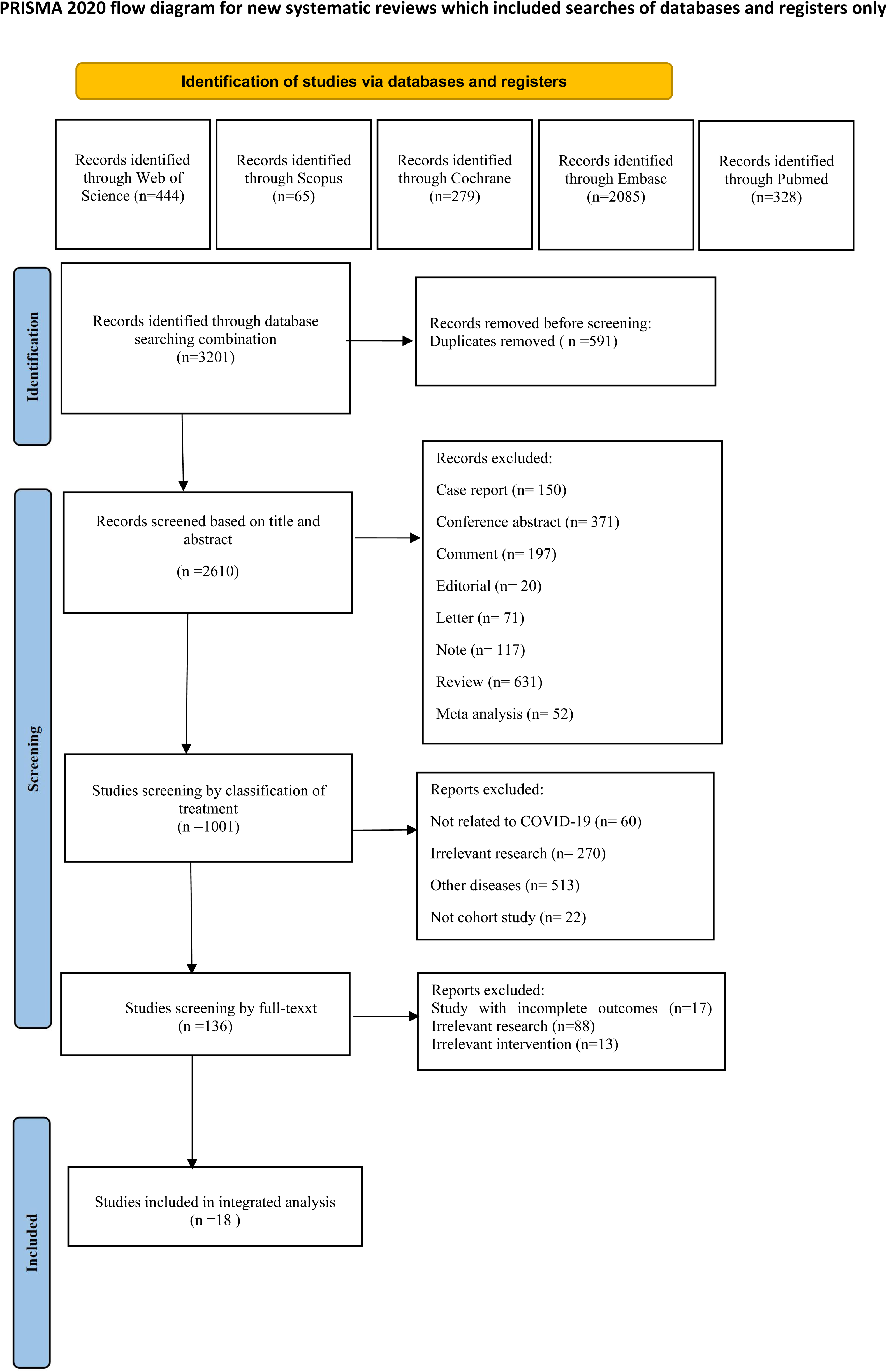
Flow diagram outlining the search strategy and study identification for meta-analysis.

The characteristics of the included studies are presented in Table. 1 Eighteen cohort studies, including 3 prospective cohort studies and 15 retrospective cohort studies, were conducted in developed and developing countries involving different races. The total number of patients included in these studies was 46, 083, 975 of which more than 37, 475, 604 were in the no COVID-19 group, and over 8, 327, 779 were in the COVID-19 group. The duration of the studies ranged from 5 to 39 months. The male ranged from 41.2% to 90.42%, and the mean age of the participants ranged from 27.99 (±7.96) to 73.85 (±4.50) years. Only seven studies reported the mean Body Mass Index (BMI) of the no COVID-19 group, which ranged from 26 (±5.0) kg/m^2^ to 30.2 (±5.7) kg/m^2^, while the mean BMI of the COVID-19 group ranged from 27.1 (±6.7) kg/m^2^ to 31.00 (±6.06) kg/m^2^. Furthermore, there were eight studies collected the smoking history of the individuals: 5.52% to 57.43% in no COVID-19 group while 4.41% to 59.69% in COVID-19 group. In addition, eleven researches provided the prevalence of diabetes: 7.42% to 77.22% in no COVID-19 group while 8.34% to 75.61% in COVID-19 group. What’s more, there were also five studies recorded the prevalence of high cholesterol: 8.63% to 74.29% in no COVID-19 group while 8.80% to 72.79% in COVID-19 group. Lastly, three articles mentioned the morbidity of coronary heart disease: 22.12% to 78.21% in no COVID-19 group, then, 22.12% to 35.74% in COVID-19 group.

**Table 1.**
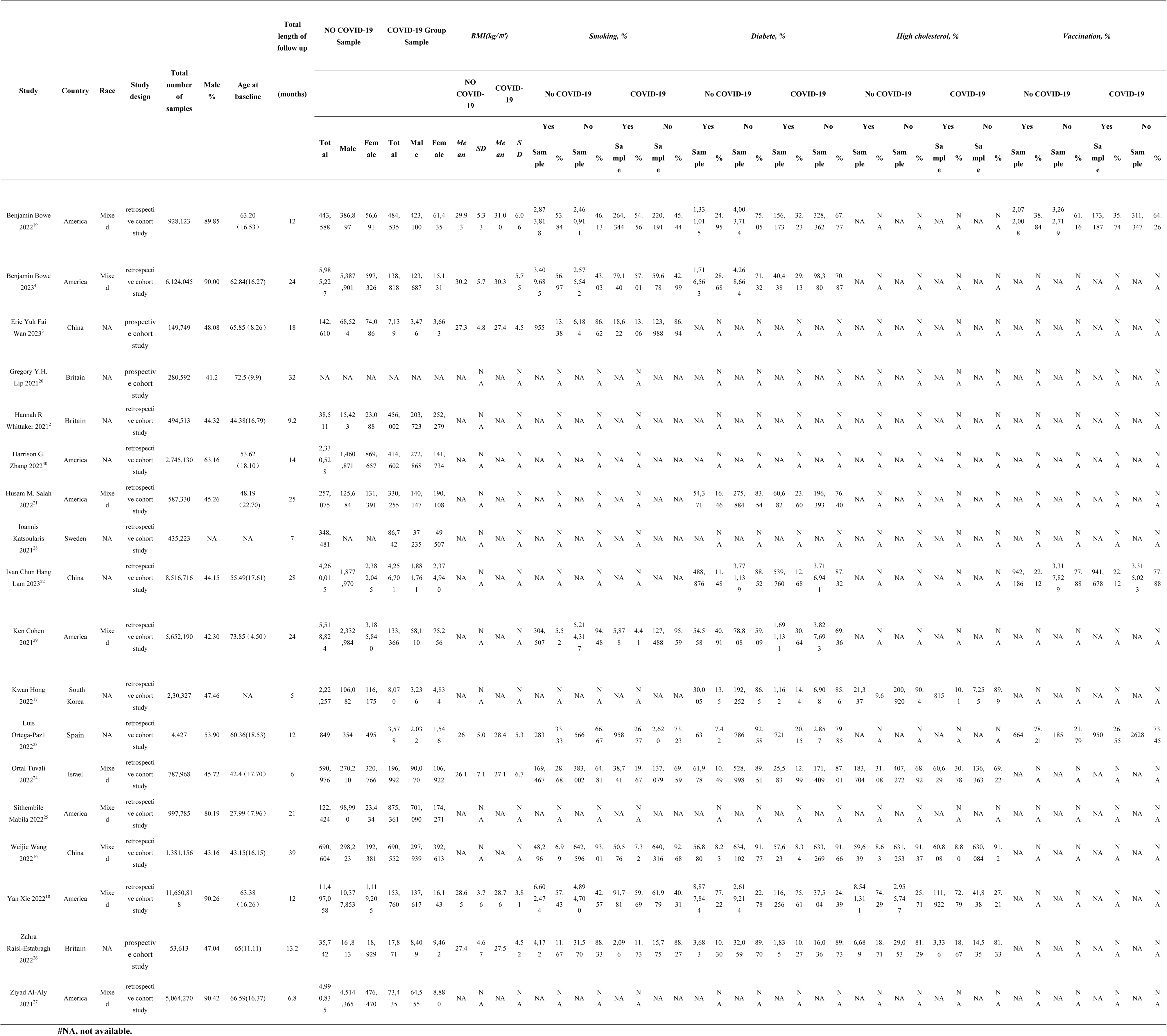
Characteristics of the included studies in the meta-analysis.

### 3.2. Quality assessment

We used the NOS, a measure of bias, to analyze the case-control and cohort studies. Only studies at least with a minimum score of 7 were deemed to be of high quality. In our meta-analysis, fifteen articles had a performance score of more than 7^2–4,16–27^, and three studies scored 6 as medium quality^28–30^, as shown in Table. 2.

**Table. 2.** Quality assessment of the included studies in the meta-analysis.

| **Source** | **Selection** | **Comparability** | **Outcome** | **Total Score** |
| --- | --- | --- | --- | --- |
| **Benjamin Bowe 2022<sup>19</sup>** | +++ | ++ | +++ | 8 |
| **Benjamin Bowe 2023<sup>4</sup>** | ++++ | ++ | +++ | 9 |
| **Eric Yuk Fai Wan 2023<sup>3</sup>** | +++ | ++ | ++ | 7 |
| **Gregory Y.H. Lip 2021<sup>20</sup>** | ++++ | ++ | ++ | 8 |
| **Hannah R Whittaker 2021<sup>2</sup>** | ++++ | ++ | ++ | 8 |
| **Harrison G. Zhang 2022<sup>30</sup>** | +++ | ++ | + | 6 |
| **Husam M. Salah 2022<sup>21</sup>** | +++ | ++ | ++ | 7 |
| **Ioannis Katsoularis 2021<sup>28</sup>** | +++ | ++ | + | 6 |
| **Ivan Chun Hang Lam 2023<sup>22</sup>** | ++++ | ++ | ++ | 8 |
| **Ken Cohen 2021<sup>29</sup>** | +++ | ++ | + | 6 |
| **Kwan Hong 2022<sup>17</sup>** | ++++ | ++ | + | 7 |
| **Luis Ortega-Pazl 2022<sup>23</sup>** | ++++ | ++ | ++ | 8 |
| **Ortal Tuvali 2022<sup>24</sup>** | ++++ | ++ | ++ | 8 |
| **Sithembile Mabila 2022<sup>25</sup>** | +++ | ++ | ++ | 7 |
| **Weijie Wang 2022<sup>16</sup>** | +++ | ++ | ++ | 7 |
| **Yan Xie 2022<sup>18</sup>** | +++ | ++ | ++ | 7 |
| **Zahra Raisi-Estabragh 2022<sup>26</sup>** | ++++ | ++ | ++ | 8 |
| **Ziyad Al-Aly 2021<sup>27</sup>** | ++++ | ++ | ++ | 8 |

#### 3. 3. 1 Cardiovascular disease risk in long COVID-19 patients

Compared to the control cohort, our meta-analysis showed a pooled OR of 1.87 (95% CI 1.56-2.25) (I^2^ = 99.4%, p= 0.000) for the overall risk of cardiovascular events, while major cardiovascular diseases (HR 1.52; 95% CI, 1.42-1.63; I^2^ = 69.1%, P= 0.021) and cardiovascular mortality (HR 1.65; 95% CI, 1.12-2.43; I^2^ = 39.7%, P= 0.173), indicating a higher risk among long COVID-19 patients (Supplementary material online, Appendix Figure. 1 A-C)

We noted a significant positive association between long COVID-19 and 17 cardiovascular disease outcomes, consisting of acute coronary disease (RR 1.44; 95% CI, 1.21-1.70; I^2^ = 87.8%, P= 0.000) (Supplementary material online, Appendix Figure. 2), angina (RR 1.33; 95% CI, 1.17-1.52; I^2^ = 80.4%, P= 0. 000) (Supplementary material online, Appendix Figure. 3A), arrhythmia ( RR 1.99; 95% CI, 1.44-2.74; I^2^ = 98.3%, P= 0.000) (Supplementary material online, Appendix Figure. 4A), atrial fibrillation (OR 1.48; 95% CI, 1.25-1.75; I^2^ = 93.3%, P= 0.000) (Supplementary material online, Appendix Figure. 5A), atrial flutter (RR 1.31; 95% CI, 0.97-1.76; I^2^ = 92.2%, P= 0.000) (Supplementary material online, Appendix Figure. 6), bradycardia (OR 1.48; 95% CI, 1.34-1.64; I^2^ = 88.2%, P= 0.000) (Supplementary material online, Appendix Figure. 7), cardiac arrest (RR 1.95; 95% CI, 1.48-2.58; I^2^ = 63.4%, P= 0.018) (Supplementary material online, Appendix Figure. 8), cardiogenic shock (RR 1.85; 95% CI, 1.59-2.14; I^2^ = 0.0%, P= 0.604) (Supplementary material online, Appendix Figure. 9), cardiomyopathy (RR 1.52; 95% CI, 1.25-1.83; I^2^ = 93.4%, P= 0.000) (Supplementary material online, Appendix Figure. 10A), coronary disease(HR 1.65; 95% CI, 1.28-2.14; I^2^ = 81.7%, P= 0.004) (Supplementary material online, Appendix Figure. 11), heart failure (OR 1.77; 95% CI, 1.52-2.05; I^2^ = 96. 4%, P= 0.000) (Supplementary material online, Appendix Figure. 12A), hypertension (HR 2.39; 95% CI, 0.60-9.62; I^2^ = 99.6%, P= 0.000) (Supplementary material online, Appendix Figure. 13), ischemic heart disease (HR 1.49; 95% CI, 1.13-1.96; I^2^ = 97.3%, P= 0.000) (Supplementary material online, Appendix Figure. 14), myocardial infarction (OR 1.63; 95% CI, 1.42-1.86; I^2^ = 80.9%, P= 0.000) (Supplementary material online, Appendix Figure. 15), myocarditis (OR 4.42; 95% CI, 2.94-6.67; I^2^ = 69.7%, P= 0.000) (Supplementary material online, Appendix Figure. 16), pericarditis (OR 2.07; 95% CI, 1.59-2.69; I^2^ = 85.9%, P= 0.000) (Supplementary material online, Appendix Figure. 17) and tachycardia (OR 1.76; 95% CI, 1.63-1.90; I^2^ = 82.8%, P= 0.000) (Supplementary material online, Appendix Figure. 18).

Long COVID-19 patients had a higher risk of developing cardiovascular disease risk in relevant studies generally, according to the combined multivariate RR of 19 outcomes. A forest plot of the OR is shown in Figure. 2. We noted a significant positive association between long COVID-19 and 19 cardiovascular disease outcomes (RR 1.68; 95% CI, 1.55-1.81; I^2^ = 69.1%, P= 0.000).

**Figure 2.**
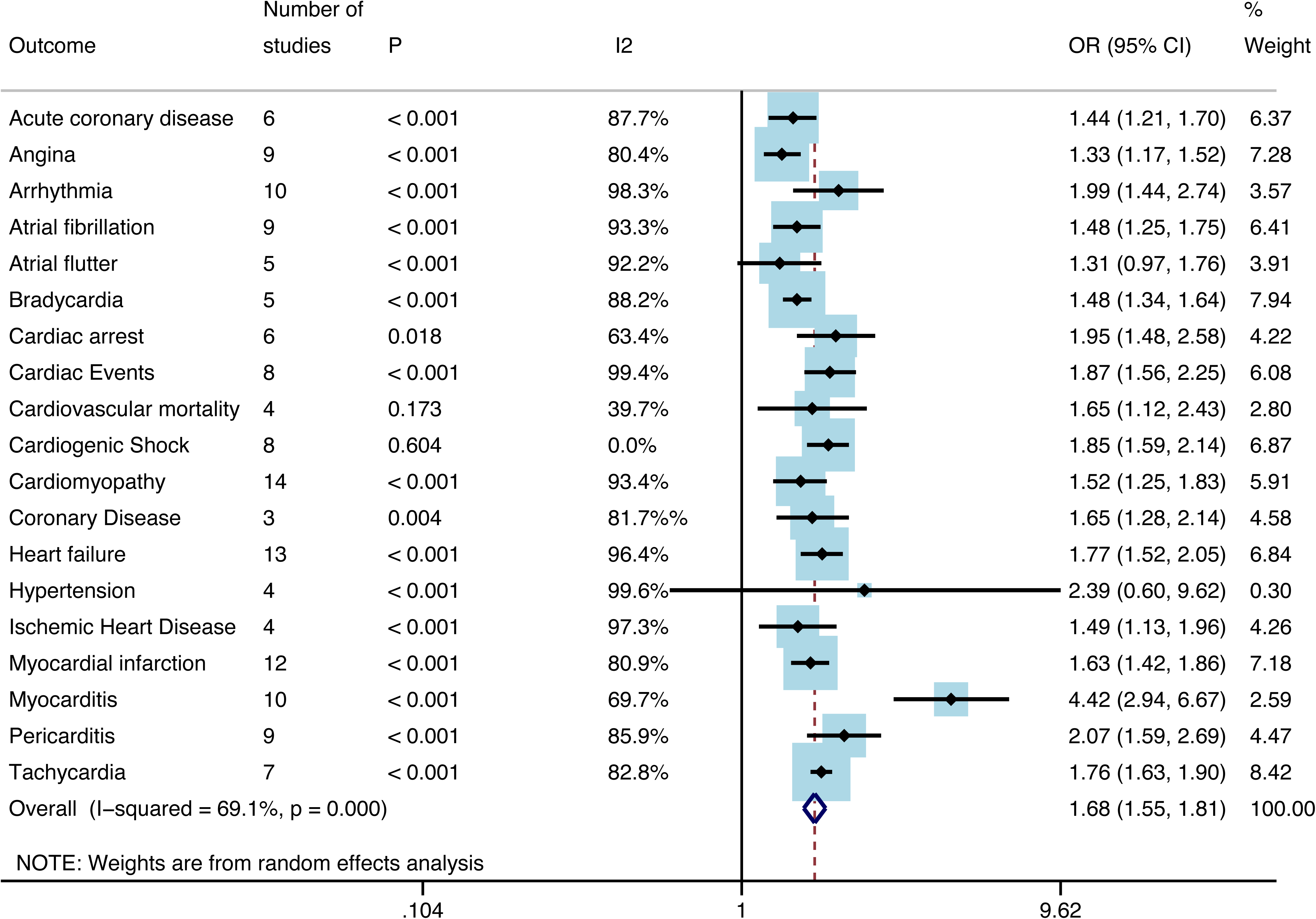
A meta-analysis of the combined multivariate relative risk of 19 outcomes.

The combined univariable OR of 4 outcomes was as well carried out, heart failure (OR 1.56; 95% CI, 0.66-3.73; I^2^ = 97. 6%, P= 0.000), myocarditis (OR 2.72; 95% CI, 0.40-18.32; I^2^ = 95.1%, P= 0.000), myocardial infarction (OR 2.37; 95% CI, 0.74-7.61; I^2^ = 77.5%, P= 0.035), pericarditis (OR 1.47; 95% CI, 0.29-7.42; I^2^ = 94. 9%, P= 0.000), suggesting that confounding factors are prevalent, potentially compromising the reliability of the results in univariate analysis, as shown in Figure. 3. However, the significance of aggregated analysis persists (OR 1.97; 95% CI, 1.11-3.52; I^2^ = 95.5%, P= 0.000).

**Figure 3.**
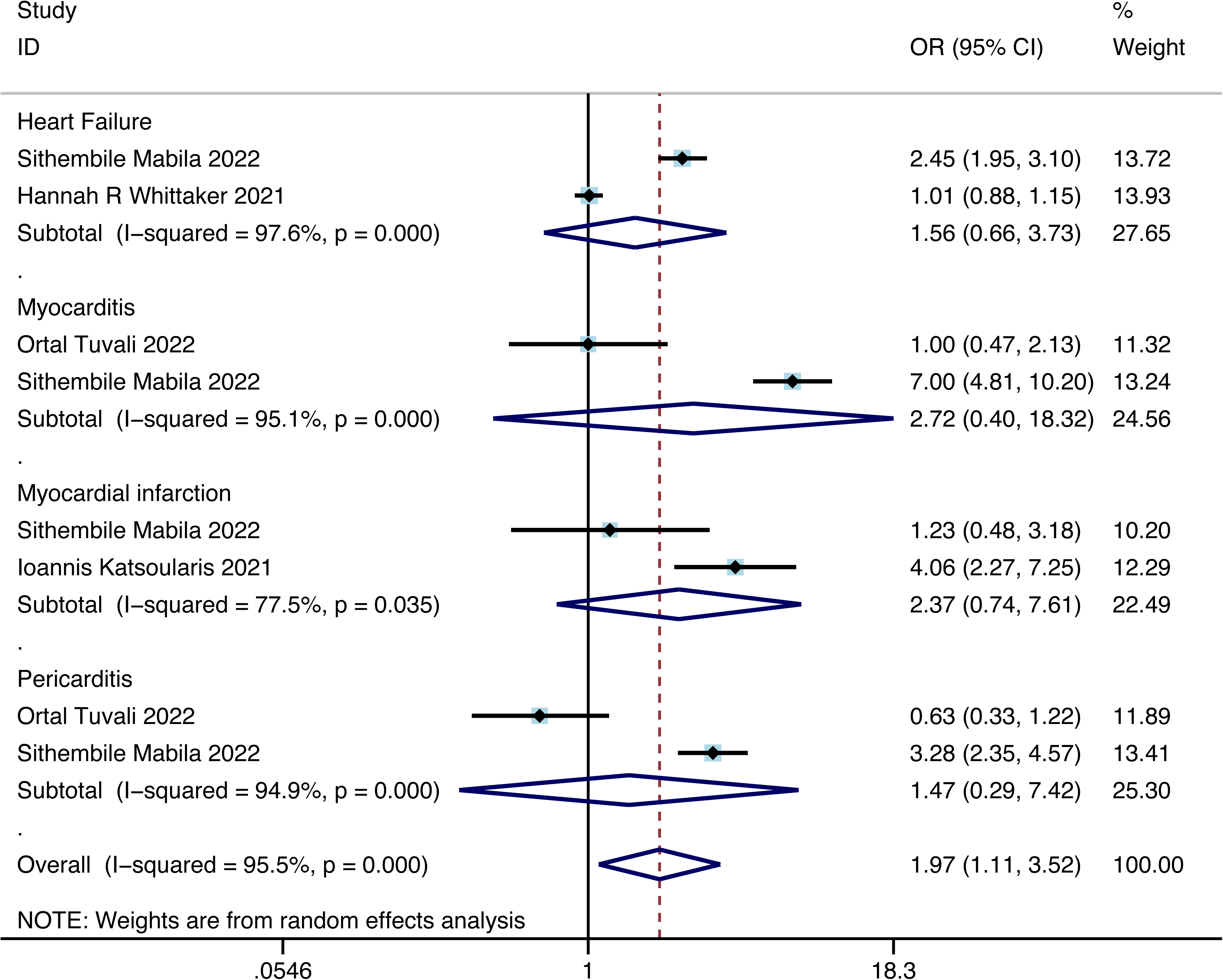
A meta-analysis of the combined univariable the combined OR of 4 outcomes.

#### 3. 3. 2 Subgroup Analysis

A subgroup analysis was performed according to type of inpatient and outpatient, for inpatient: angina (RR 1.70; 95% CI, 0.97-2.99; I^2^ = 87.9%, P= 0.004) and heart failure (RR 1.47; 95% CI, 1.21-1.79; I^2^ = 86.0%, P= 0.000), as shown in Supplementary material online, Appendix Figure. 3B and 12B, respectively; for outpatient atrial fibrillation and flutter ( RR 1.11; 95% CI, 0.82-1.52; I^2^ = 87.4%, P= 0.005) and heart failure ( RR 1.06; 95% CI, 0.95-1.17; I^2^ = 0.0%, P= 0.879), as shown in Supplementary material online, Appendix Figure. 5B and 12B, respectively. Judging from the outcomes, it showed that the risk of HF significantly increased in the inpatient group, may reflect severe COVID-19 would increase the risk of long-term HF to some extent. Furthermore, there were 2 main subgroups for the pathological type of cardiomyopathy, the subgroup analysis of ischemic cardiomyopathy (RR 1.58; 95% CI, 1.11-2.26; I^2^ = 92.7%, P= 0.000) and non-ischemic cardiomyopathy (RR 1.34; 95% CI, 1.10-1.64; I^2^ = 78.8%, P= 0.000), as shown in Supplementary material online, Appendix Figure. 10B.

Meanwhile, there were also two other subgroup analyses, comprising ventricular arrhythmias (RR 1.70; 95% CI, 1.36-2.13; I^2^ = 84.9%, P= 0.000) and atrial fibrillation and flutter (RR 1.78; 95% CI, 0.97-3.25; I^2^ =98.5%, P= 0.000), as shown in Supplementary material online, Appendix Figure. 4B and 5C.

Considering the limitation of retrospective cohort study, we also conducted the risk of cardiovascular outcomes only based on prospective cohort studies, which is more reliable. It displayed that atrial fibrillation (HR 1.60; 95% CI, 0.87-2.95; I^2^ = 94.6%, P= 0.000), heart failure (HR 1.74; 95% CI, 1.25-2.41; I^2^ = 65.9%, P= 0.053), pericarditis (HR 4.47; 95% CI, 3.15-6.35; I^2^ = 0.0%, P= 0.996), as shown in Supplementary material online, Appendix Figure. 19. Also Pooled analysis demonstrates the significance of the aforementioned three outcomes (HR 2.14; 95% CI, 1.52-3.01; I^2^ = 89.6%, P= 0.000).

### 3.4. Heterogeneity and publication bias

The heterogeneity was relatively high, but with the help of Meta-regression, we figured out that country, study design, total number of samples, age at baseline and total months of follow-up should be responsible for sources of heterogeneity to a certain extent Supplementary material online, Appendix Figure. 20-39). Meanwhile, owing to the observational research’s character, all studies inherently entail an inevitable risk of bias, irrespective of their quality rating^31^. The results of the funnel plot, Begg’s, and Egger’s tests collectively indicated the absence of publication bias in the meta-analysis basically (Supplementary material online, Appendix Figure. 40, Supplementary material online, Appendix Table. 2-19).

### 3.5. Sensitivity analysis

Regarding the results displayed above, there was no discernible alteration in the stability of the meta-analysis (Supplementary material online, Appendix Figure. 41).

## 4. Discussion

At present, a definitive mechanism explaining the pathophysiology of long-COVID contributing to cardiovascular diseases remains not conclusive. One hypothesis proposes that myocardial injury in COVID-19 may arise from either direct viral invasion of cardiomyocytes or the presence of circulating inflammatory mediators, including cytokines and/or endotoxins, or potentially a combination of both factors^32^. Upon infection, there was an observed upregulation of ACE2, the receptor for SARS-CoV-2, in the pulmonary alveolar epithelial barrier, vascular endothelial cells, and cardiomyocytes^33^. In single-nuclei RNA sequencing studies, robust expression of ACE2 in the heart is evident in pericytes, vascular smooth muscle cells, fibroblasts, and cardiomyocytes^34^. An alternative hypothesis suggests that both hypo- and hyper-immune responses may play a role in the severity of COVID-19, potentially leading to systemic inflammatory responses or a cytokine storm, culminating in cell death and multi-organ dysfunction^35^. According to cell and animal models of SARS-CoV-2 infection, coupled with transcriptional and serum profiling of individuals with COVID-19, one experiment revealed a consistent and distinctive inflammatory reaction, whose response was characterized by reduced levels of type I and III interferons concomitant with elevated chemokine levels and prominent expression of Interleukin-6 (IL-6)^36^.

Numerous cohort studies have identified an association between long-term COVID-19 and diverse cardiovascular outcomes. However, a comprehensive systematic meta-analysis assessing composite cardiac outcomes is currently lacking. This study employed meta-analysis techniques to evaluate the relationship between long COVID-19 and cardiovascular outcomes. Our meta-analysis, based on the scrutiny of 18 articles comprising both retrospective and prospective studies, encompassing 46, 083, 975 participants, revealed that individuals with long COVID-19 face an increased risk of developing cardiovascular diseases overall. Although the relatively high heterogeneity is acknowledged, it is inevitable for the diversity arising from variations in countries, study designs, total sample sizes, baseline ages, and durations of follow-up, as revealed by the results of meta-regression. Additionally, the complexity of confounding factors presents a challenge in achieving complete standardization. All included studies, even those with a high rating, inherently entail an unavoidable risk of bias, owing to their observational characteristic^31^. And we also hope to collect the data of long Covid patients’ BMI^37^, smoking status^38–40^, diabetes^41^, high cholesterol^42,43^, vaccination^44–46^ etc as much as possible, further identifying sources of heterogeneity. But in our study, all the HRs, RRs and ORs are all over >1.0, as well as the absence of publication bias based on the results of the funnel plot, Begg’s, and Egger’s tests, suggesting the model is robust and reliable.

Hence, we propose that routine assessment of cardiac function is imperative for the population with long COVID-19 who are at a heightened risk of cardiovascular disease. Timely interventions have the potential to mitigate the risk of cardiovascular diseases and associated mortality. At present, the management of long COVID and CV sequelae encompass a range of therapeutic interventions^5^: these interventions comprise various rehabilitation programs, both through telemedicine and in-person consultations, targeting the amelioration of symptoms such as fatigue, cognitive decline, and breathlessness; additionally, immunomodulatory treatments, including steroids, laranilubmab, tocilizumab, atorvastatin, and colchicine, are also under scrutiny; meanwhile, antifibrotic agents, like pirfenidone and LYT-100, as well as anticoagulation strategies employing apixaban, are being experimented; furthermore, there are therapies directed towards cognition enhancement, such as transcranial stimulation, as well as metabolic modulators like niagen. What’s more, Paxlovid demonstrates higher efficacy in those harboring preexisting neurological or cardiovascular conditions, individuals with immunosuppression and older patients^47^. But, the preeminent strategy for averting severe complications stemming from SARS-CoV-2 infection is still vaccination^48,49^. Furthermore, vigilant monitoring of viral infection-associated cardiac injury remains imperative throughout the management of COVID-19.

## 5. Limitations

We acknowledge certain limitations in our study that warrant consideration. Firstly, several of our conducted meta-analyses displayed notable statistical heterogeneity, which may potentially lead to an overestimation of the strength of associations, as suggested by large meta-epidemiologic studies. The observed heterogeneity may be attributed to adjusted confounding factors and other variables within the included studies. Additionally, among the 18 articles analyzed, special attention should be given to the cumulative duration of follow-up, given the chronic nature of SARS-CoV-2 infection. Furthermore, retrospective cohort studies inherently introduce a risk of bias. Therefore, there is an ongoing need for additional relevant cohort studies, particularly prospective cohort studies, to enable a more comprehensive analysis. But it is crucial to emphasize that all studies included in our analysis provided risk factors through multivariable regression, ensuring the derivation of robust conclusions.

## 6. Conclusions

In summary, our study furnishes evidence indicating that individuals long COVID-19 face a markedly elevated risk of developing cardiovascular outcomes. Implementing intensified vaccination strategies, particularly with the latest COVID-19 vaccine, in populations at a severe risk, and regularly monitoring cardiac function in those at a high cardiovascular risk, may prove beneficial in mitigating the risk of cardiovascular diseases and associated mortality in individuals with long COVID-19.

## Data Availability

All data produced are available online at

## Abbreviations

WHO: World Health Organization;
HR: Hazard ratio;
RR: risk ratio;
OR: odds ratio;
CI: confidence interval;
REM: random-effects model;
ACE2: angiotensin-converting enzyme 2;
SARS-CoV-2: Severe Acute Respiratory Syndrome Coronavirus 2 CoV;
MESH: Medical Subject Headings;
NOS: Newcastle-Ottawa Scale;
FEM: fixed-effect model;
BMI: Body Mass Index;
IL: interleukin;

## Author Contributions

Study design: ZC, YH; Data collection: YH, ZW, HL, YL; Data analyses: YH, ZC; Results visualization: YH; Results interpretations: YH, ZC; Manuscript writing: YH, ZC; Manuscript revising: YH, ZW, HL, YL ZC. All authors approved the final version of the manuscript.

## Data Availability

The research contributions presented in this article are detailed in the article/supplementary material; you can contact the relevant author for further information.

## Declarations

Competing interests The authors declare that they have no competing interests.

